# The diabetic myocardial transcriptome reveals Erbb3 as a novel biomarker of incident heart failure

**DOI:** 10.1101/2024.01.05.24300893

**Authors:** Marcella S Conning-Rowland, Marilena Giannoudi, Michael Drozd, Oliver I Brown, Nadira Y Yuldasheva, Chew W Cheng, Paul J Meakin, Sam Straw, John Gierula, Ramzi A Ajjan, Mark T Kearney, Eylem Levelt, Lee D Roberts, Kathryn J Griffin, Richard M Cubbon

## Abstract

**Aims:** Diabetes mellitus (DM) increases heart failure incidence and worsens prognosis, but the molecular basis of diabetic heart disease is poorly defined in humans. We aimed to define the diabetic myocardial transcriptome and validate hits in their circulating protein form to define disease mechanisms and biomarkers.

**Methods and Results:** RNA-sequencing data from the Genotype-Tissue Expression (GTEx) project was used to define differentially expressed genes (DEGs) in right atrial (RA) and left ventricular (LV) myocardium from people with versus without DM (type 1 or 2). DEGs were validated in their circulating protein form in the UK Biobank cohort, searching for directionally concordant differential expression. Validated plasma proteins were characterized in UK Biobank participants, irrespective of diabetes status, using cardiac magnetic resonance imaging, incident heart failure and cardiovascular mortality.

We found 32 and 32 DEGs associated with DM in the RA and LV, respectively, with no overlap between these. Plasma proteomic data was available for 6 hits, with only ERBB3 (LV hit) exhibiting directional concordance, being lower in myocardium and blood of people with DM. Irrespective of DM status, lower circulating ERBB3 was associated with impaired left ventricular contractility and higher LV mass. Participants in the lowest quartile of circulating ERBB3 had increased incident heart failure and cardiovascular death vs. participants in all other quartiles. Similar associations were noted for ERBB2 and ERBB4.

**Conclusions:** DM is characterized by lower ERBB3 expression in the myocardium and lower plasma protein concentration. This is associated with left ventricular dysfunction, incident heart failure and cardiovascular mortality.

## 1. Introduction

The global prevalence of diabetes mellitus (DM) was estimated at 9.3% of adults (463 million people) in 2019 and is expected to rise to 10.9% (700 million people) by 2045.^1^ Whilst the incidence of most microvascular and macrovascular complications of DM continues to decline, the incidence of heart failure (HF) is static.^2^ Epidemiological studies suggest that people with DM experience at least a doubled incidence of HF, both for those with type 1 or type 2 DM, with comparable incidence rates for HF with preserved or reduced ejection fraction.^3,4^ Moreover, the prognosis of people with HF who also have DM is worse, with doubled rates of all-cause mortality^5^, and 50% greater rates of HF hospitalisation, compared to individuals with normal glucose metabolism.^6^ Whilst obesity is an important risk factor for HF in people with DM, it is notable that even those people without obesity or other modifiable risk factors experience increased risk of HF and myocardial dysfunction.^2,7^ This suggests that existing risk factor modification strategies are not fully addressing the pathophysiology of diabetic heart disease.

Our understanding of the molecular basis of myocardial disease associated with DM largely comes from animal models that do not recapitulate many of the phenotypic and treatment characteristics of people with DM.^8–10,11^ This reflects the challenges of collecting myocardial biopsies, especially of the left ventricle, from large cohorts of people with and without DM. Moreover, many studies apply a biased approach to molecular characterisation, focusing on predefined genes or pathways, which further increases the risk of missing unappreciated pathophysiological processes. We aimed to address this by using RNA-sequencing (RNA-seq) data from the Genotype-Tissue Expression (GTEx) project, which collected multi-organ biopsy material from a large cohort of organ donors.^12^ Differentially expressed genes (DEGs) associated with DM were defined separately in right atrial (RA) and left ventricular (LV) tissue. These were further explored using plasma proteomic data from the UK Biobank (UKB) cohort to seek concordant hits with potential use as biomarkers and to inform understanding of pathophysiology.

## 2. Methods

### 2.1 Myocardial RNA-seq data from GTEx project

The GTEx project was established by the Broad Institute of MIT and Harvard with a primary goal of defining associations between genomic variation and gene expression in 54 tissues across the human body.^12^ Tissues were retrieved post-mortem from organ donors following consent from family-decision makers. RNA-seq was performed as described in detail on the GTEx project website (https://gtexportal.org/home/methods). RA appendage and LV apex RNA-seq data are available from 429 and 432 donors, although our analyses pertain to 425 RA and 428 LV samples after exclusion due to missing metadata required as covariates in differential gene expression analysis. Bulk RNA-seq raw count data were downloaded directly from the GTEx portal (https://www.gtexportal.org/home/), using GTEx version 8 release on 18^th^ July 2022. This includes metadata on donor age, sex, ischemic time (between death and sample collection) and Hardy score (death classification as: 0) Ventilator case for instances of death on a ventilator immediately before death; 1) Violent and fast death [e.g. accident, blunt force trauma or suicide] with terminal phase estimated at <10min; 2) Fast death of natural causes [e.g. sudden unexpected deaths of people who had been reasonably healthy] after a terminal phase estimated at <1hr; 3) Intermediate death after a terminal phase of 1-24 hrs in patients who were ill but death was unexpected; 4) Slow death after a long illness, with a terminal phase longer than 1 day. For sensitive metadata, pertaining to comorbidities, a protected access data application was granted by dbGaP (https://www.ncbi.nlm.nih.gov/gap/) #32524 "Defining tissue-specific transcriptional profiles associated with diabetes".

### 2.2 RNA-seq analysis pipeline for GTEx data

Differential Gene Expression analysis was conducted using R (Version 4.1.1) and the DESeq2 package (Version 1.36.0).^13^ We sought differentially expressed genes (DEGs) associated with diabetes. Donors with type 1 or type 2 diabetes were pooled to create a single diabetes variable, since multiple donors were recorded as having both forms of diabetes. Those with diabetes status unknown were excluded. Covariates in DESeq2 analyses were: age (20-29, 30-39, 40-49, 50-59, 60-69, 70-79); sex; race; ischemic time as categories of 300 minutes (0-299, 300-599, 600-899, 900-1199, 1200-1499); Hardy score; BMI which was classed as normal weight (18.5–24.9[kg/m^2^), overweight (25–29.9[kg/m^2^), and obese (>30 kg/m^2^); medical history indication of hypertension and myocardial infarction; and RNA integrity score (RIN)(5.1-6, 6.1-7, 7.1-8, 8.1-9, 9.1-10). Technical factors including ischemic time, defined by GTEx as “time from death or withdrawal of life-support until the time the sample is placed in a fixative solution or frozen”, Hardy Score and RIN were included as these either directly related to, or had the possibility to alter RNA quality and/or expression. Biological cofactors of age, sex, race and BMI were included because these are also known to alter cardiac gene expression. Genes were filtered to include only those with greater than 10 read counts in at least the same number of samples as included in the diabetes subgroup. Effect size shrinkage using the *apeglm* method was applied for visualisation and ranking of genes.^14^ False discovery rate (FDR) adjusted p values produced by DESeq2 using the Benjamini-Hochberg method were used, with adjusted p<0.05 defined as statistically significant. Functional profiling of DEGs was performed using gProfiler (https://biit.cs.ut.ee/gprofiler/gost). DEGs were run as queries, with significance threshold calculated using Benjamini-Hochberg FDR. Driver terms were highlighted in the output as the most relevant Gene Ontology terms and exported.

### 2.3 UK Biobank cohort

UKB is a prospective observational cohort study of 502,462 participants aged 37-73 years, recruited from 22 assessment centres across the United Kingdom (UK) between 2006-10. It is an open access resource developed using UK Government and biomedical research charity funding which linked wide-ranging phenotypic and health care record data. The UK Biobank resource is open to all bona fide researchers. Full details of its design and conduct are available online (https://www.ukbiobank.ac.uk). UKB received ethical approval from the NHS Research Ethics Service (11/NW/0382); we conducted this analysis under application number 105351. All participants provided written informed consent and the research was conducted in line with the Declaration of Helsinki. All analyses were conducted via the UKB Research Analysis Platform (https://ukbiobank.dnanexus.com).

### 2.4 Definition of diabetes in UKB

Baseline sociodemographic characteristics and comorbidities were recorded by participants completing a touchscreen and nurse-led interview at study recruitment and used as we have previously described.^15^ Data from face-to-face nurse-led interview was used to ascertain baseline comorbidities and medication. Diabetes was classified as any of “diabetes” (UK Biobank field ID ‘1220’); “type 1 diabetes mellitus” (‘1222’); “type 2 diabetes mellitus” (‘1223’); “diabetic eye disease” (‘1276’); “diabetic neuropathy/ulcers” (‘1468’) and “diabetic nephropathy” (‘1607’), as we have previously described.^7^

### 2.5 Plasma proteomic data in UKB

A recent update to UKB is the addition of plasma proteomic profiles from 54,219 UKB participants.^16^ At time of analysis, data were available for 1536 proteins, measured on Olink Explore 1536 panel, a proximity extension assay using paired antibodies and complimentary oligonucleotides. This panel includes proteins grouped into categories of proteins related to; oncology, inflammation, neurology and cardiometabolic related proteins. The cardiometabolic protein set includes clinically relevant biomarkers of cardiac stress and/or injury including NT-proBNP and TNNI3 (Troponin I). Details of UKB quality control procedures for plasma proteomics have been descried by Sun and colleagues.^16^ Protein concentration is provided as normalized protein expression (NPX) values. Where proteins were below the limit of assay detection in specific samples, these are recorded as missing data. NPX values were converted to z scores, Z score was defined as z = (x-μ)/σ, where x = protein NPX, μ = population mean, and σ = population standard deviation. We used proteomic data collected at the initial assessment centre visit.

### 2.6 Cardiovascular Magnetic Resonance Imaging data in UKB

From 2014, approximately 50,000 participants have taken part in multimodality imaging assessment, which included cardiovascular magnetic resonance imaging (MRI).^17–19^ Expression of ERBB3 was compared against MRI parameters to determine if protein expression related to MRI markers of cardiac function. MRI parameters used were: left ventricular ejection fraction (LVEF) (UKB Field ID: ‘22420’), LV circumferential strain global (UKB Field ID: ‘24157’), LV longitudinal strain global (‘24181’), LV radial strain global (‘24174’), LV myocardial mass (‘24105’), myocardial wall thickness global (‘24140’). Of the UKB participants who had ERBB3 protein expression data, there was LV imaging data on approximately 5000 of these (ranging from 4,986 participants for LV longitudinal strain global to 5,122 participants for LV myocardial mass). Additionally, cardiac contractility index (CCI) was used as an additional measure of cardiac functionality. CCI was derived by systolic blood pressure (UKB field ‘4080’) divided by LV end-systolic volume index, calculated as LV end systolic volume (LVESV) (‘22422’) normalised to body surface area (‘22427’) as previously described.^7^ When calculating CCI, 6 participants with LVESV <20ML were excluded as outliers of >3 standard deviations from the mean.

### 2.7 Definition of outcome measures in UKB

Time to heart failure was defined as the time between date of baseline assessment centre visit (when plasma proteins were measured) and the date that heart failure first reported (UKB field IDs 131354 and 131355), excluding participants with pre-existing heart failure. Self-reported heart failure codes were excluded (UKB Code IDs 50 and 51 in data field 131355) to focus solely on events confirmed in medical records, including the death register, primary care, and hospital admission reports (UKB Code IDs 20, 21, 30, 31, 40, 41 in data field 131355). Cardiovascular mortality was defined I00-I99, excluding those related to infection mortality, as previously described.^7^

### 2.8 Statistical analysis of UKB

Categorical data are presented as number (percentage of denominator) and continuous data are presented as mean (standard deviation). Statistical significance was defined as p<0.05 using 2-sided tests. Normality was determined by skewness and kurtosis tests using the Moments (v0.14.1) R package (https://cran.r-project.org/web/packages/moments/index.html). Correlations between ERBB3 and other continuous variables were calculated using Spearman’s Rank test. Comparisons across ERBB3 quartile groups were made using Chi-squared tests followed by pairwise proportional tests for categorical data, and one-way ANOVA followed by Tukey post-hoc tests for continuous data. Where a one-way ANOVA was not appropriate due to non-normality of data, Welch’s ANOVA followed by Games-Howell post-hoc tests were used as an alternative. Time to event analyses were performed using the R package Survival (v3.2.13), ^20^. The censorship date for these analyses was Aug 30^th^ 2023. As the proportional hazard assumptions were not met by Cox regression models, Poisson regression models including exposure time were generated instead.

## 3. Results

Bulk RNA-seq data from GTEx was used to identify differentially expressed genes (DEGs) in samples from people with DM versus people without DM. Of RA donors 23.5% had DM, whilst this was 23.3% for LV donors. Overall, there was a greater proportion of men than women, and the majority of donors were white. There was a greater prevalence of prior myocardial infarction and hypertension in the DM donors (**Table 1**). BMI, ischemic time, and Hardy score was comparable in donors with and without DM. As all covariates in **Table 1** have important implications for gene expression in individual samples, they were included as covariates in differential gene expression analyses, even if data were similar in groups with and without DM.

**Table 1:** Characteristics of donors with diabetes in the GTEx cohort.

|  | Right Atrium |  | Left Ventricle |  |
| --- | --- | --- | --- | --- |
|  | Non-DM | DM | Non-DM | DM |
|  | (n=305) | (n=120) | (n=329) | (n=100) |
| **Male** | 201 (65.9) | 89 (74.2) | 215 (65.3) | 76 (76) |
| **Race:** |  |  |  |  |
| **American Indian or Alaskan Native** | 0 (0) | 1 (0.8) | 0 (0) | 1 (1) |
| **Asian** | 4 (1.3) | 1 (0.8) | 6 (1.8) | 1 (1) |
| **Black or African American** | 32 (10.5) | 21 (17.5) | 35 (10.6) | 18 (18) |
| **White** | 267 (87.5) | 97 (80.8) | 286 (86.9) | 80 (80) |
| **Unknown** | 2 (0.1) | 0 (0) | 2 (0.6) | 0 (0) |
| **BMI (kg/m<sup>2</sup>)** | 27.2 (4.2) | 28.3 (4.1) | 27.1 (3.9) | 28.1 (4.3) |
| **Ischemic time (mins)** | 518 (394) | 598 (419) | 450 (392) | 497 (435) |
| **History of myocardial infarction** | 42 (13.8) | 32 (26.7) | 41 (12.5) | 23 (23) |
| **History of hypertension** | 159 (52.1) | 105 (87.5) | 160 (48.6) | 84 (84) |
| **Hardy Score:** |  |  |  |  |
| **0** | 151 (49.5) | 50 (41.7) | 186 (56.5) | 57 (57) |
| **1** | 16 (5.2) | 1 (0.8) | 19 (5.8) | 1 (1) |
| **2** | 89 (29.2) | 42 (35) | 75 (22.8) | 28 (28) |
| **3** | 16 (5.2) | 10 (8.3) | 18 (5.5) | 6 (6) |
| **4** | 33 (10.8) | 17 (14.2) | 30 (9.1) | 8 (8) |

Categorical data are presented as number (%) and continuous data as mean (standard deviation)

### 3.1 DEGs associated with diabetes in the LV and RA

After accounting for confounding covariates, we identified 46 and 72 DEGs associated with DM (FDR-adjusted p<0.05) in the LV and RA, respectively (**Figure 1** and **Supplementary Tables 1-2**. Of these, 32 LV and 32 RA DEGs had a log_2_ fold change >0.32 or <-0.32 (corresponding to a 25% difference in gene expression). Notably, there was no overlap in DEGs in the LV and RA, suggesting that DM has different pathophysiological implications in ventricular versus atrial myocardium. To explore the biological themes within DEGs, functional enrichment analysis was performed using g:Profiler.^21^ In RA, this detected enrichment of immune system terms including IgG and IgA immunoglobulin complexes (GO Terms: GO:0019814, GO:0071735, GO:0071745). In LV IgG immunoglobulin complex (GO:0071735) was also altered, but the several altered LV gene ontology terms related to neuronal activity such as neuroligin family binding protein and structural constituent of myelin sheath (GO:0097109, GO:0019911) (**Supplemental Tables 3-4**).

**Figure 1:**
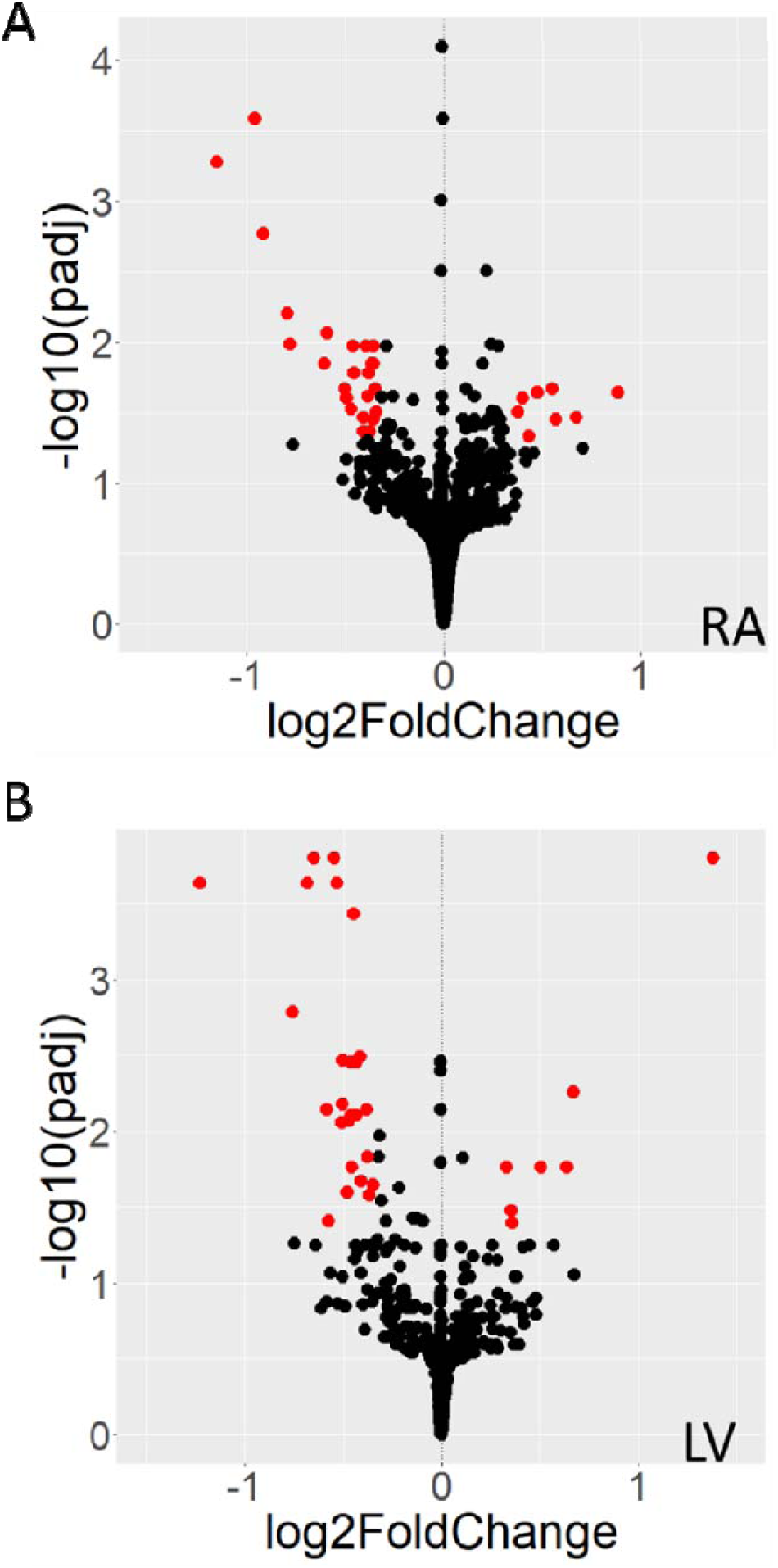
Differential gene expression associated with diabetes in the right atrium and left ventricle. Volcano plots illustrating differential gene expression for DM versus without DM for right atrium (**A**) and left ventricle (**B**). Each dot represents a gene, with red colour denoting those that achieve Benjamini-Hochberg false discovery rate-adjusted p values <0.05 and log_2_ fold change >0.32 or <-0.32 (corresponding to a 25% difference in gene expression). Raw data are presented in Supplemental Tables 1 and 2. LV – left ventricle; RA – right atrium.

### 3.2 Validation of DEGs using UKB plasma proteomics

To corroborate the DEGs observed in GTEx, we used the orthogonal approach of defining these genes at their protein level in the circulation, which may also aid the translation of these findings to clinically relevant biomarkers. UKB used Olink technology to measure 1,459 proteins in the plasma of 52,705 participants. Of the 64 DEGs observed in either LV or RA, proteomic data was available for 6: SLAMF7, MZB1, CGA, ERBB3, L1CAM, SLITRK2 (**Table 2**). All of these exhibited statistically significant differences in plasma protein expression between DM and non-DM participants. However, only ERBB3 exhibited a directionally concordant change in myocardial RNA expression and plasma protein concentration, both being lower in people with DM (**Table 2** and **Supplemental Table 5**). These data suggest that plasma ERBB3 warrants further exploration as a biomarker of cardiac disease associated with DM.

**Table 2:**
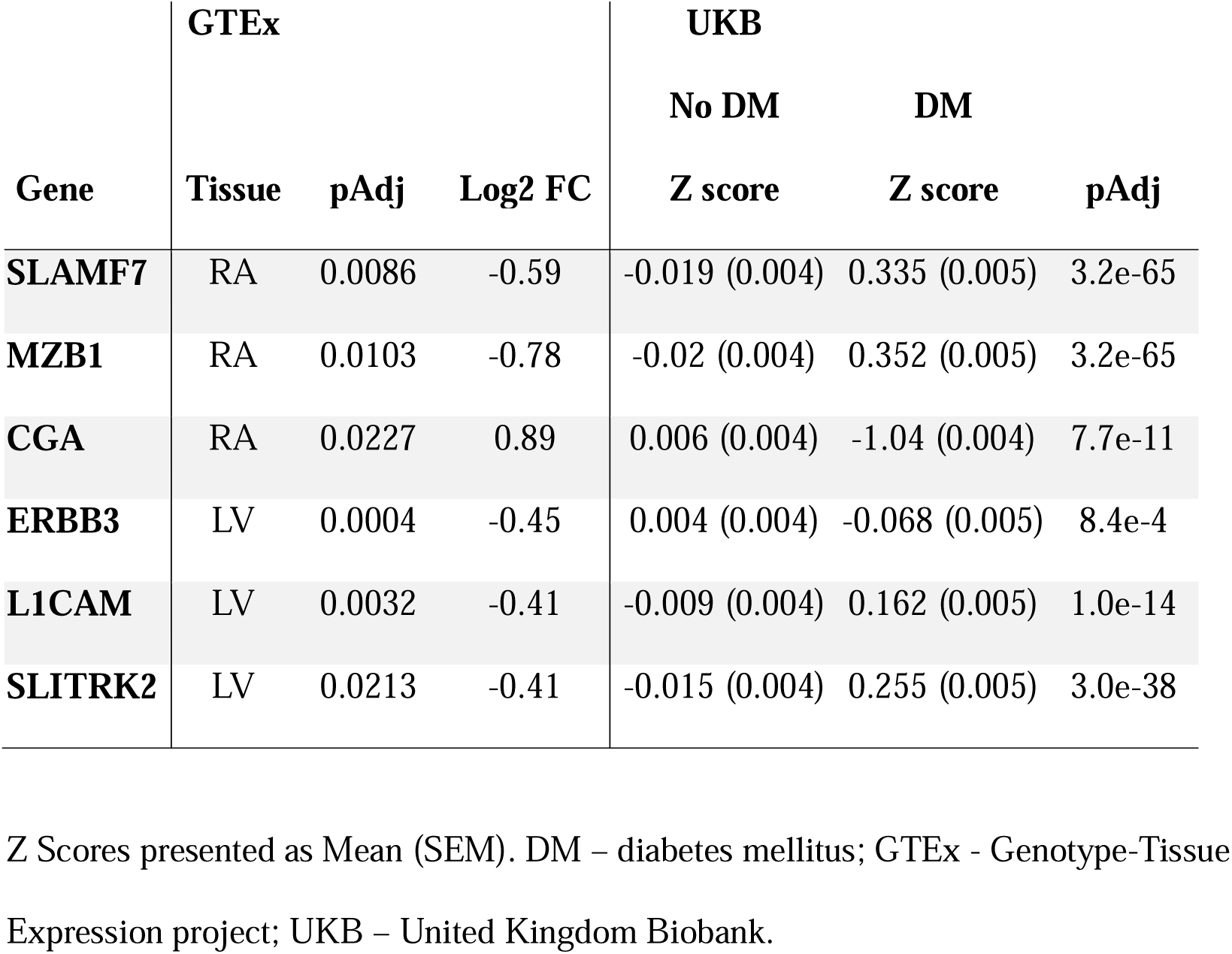
Validation of GTEx myocardial RNA-seq hits using UKB plasma proteomics.

### 3.3 Baseline characteristics of ERBB3 quartiles in UKB

To examine the wider characteristics of participants with lower circulating ERBB3 concentrations, we divided UKB participants into quartiles, with quartile 1 (Q1) having the lowest concentration of ERBB3 plasma protein (with z scores of below -0.628 for Q1, -0.628 to -0.013 for Q2, -0.013 to 0.595 for Q3 and above 0.595 for Q4). As shown in **Table 3**, age was higher in Q1 than Q4 lower (p<0.0001 across quartiles), whilst body mass index was lower (p<0.0001 across quartiles). There was also a marked male preponderance in Q1 versus Q4 (p<0.0001 across quartiles), along with a higher prevalence of DM (p<0.0001 across quartiles).

**Table 3:** Baseline characteristics of plasma ERBB3 quartiles.

|  | **Q1**<br>**(Low)** | **Q2** | **Q3** | **Q4**<br>**(High)** | **P value** |
| --- | --- | --- | --- | --- | --- |
| **Age (years)** | 57.8<br>(0.08) | 56.5<br>(0.07) | 56.4<br>(0.07) | 56.7<br>(0.07) | <0.0001 |
| **Male (%)** | 67.2<br>(8618) | 49.3<br>(6329) | 38.2<br>(4912) | 29.2<br>(3756) | <0.0001 |
| **BMI (kg/m<sup>2</sup>)** | 27.3<br>(0.04) | 27.3<br>(0.04) | 27.5<br>(0.04) | 27.9<br>(0.04) | 0.002 |
| **DM (%)** | 6.8<br>(869) | 5.0<br>(643) | 4.5<br>(579) | 5.6<br>(724) | <0.0001 |
| **Prior MI (%)** | 4.6<br>(591) | 2.4<br>(316) | 1.9<br>(243) | 1.7<br>(213) | <0.0001 |
| **Prior HF (%)** | 0.9<br>(102) | 0.5<br>(69) | 0.3<br>(45) | 0.4<br>(47) | <0.0001 |
| **Systolic BP**<br>**(mmHg)** | 136.4<br>(0.2) | 136.8<br>(0.2) | 137.9<br>(0.2) | 139.8<br>(0.2) | <0.0001 |
| **Diastolic BP**<br>**(mmHg)** | 80.6<br>(0.1) | 81.6<br>(0.1) | 82.5<br>(0.1) | 83.7<br>(0.1) | 0.002 |

Data presented as mean (SEM) for continuous data and % (n) for categorical data. ERBBB3 quartiles were defined as Z scores of: below -0.628 for Q1, -0.628 to -0.013 for Q2, -0.013 to 0.595 for Q3 and above 0.595 for Q4; each quartile includes 12,833 participants. Hospital admissions reports only of heart failure were used. BP = Blood Pressure.

To assess whether plasma ERBB3 concentration was associated with baseline cardiac function, we defined its correlation with cardiac MRI metrics, focusing on measures of LV function given ERBB3 was differentially expressed in LV myocardium (**Table 4**). Lower ERBB3 was associated with lower ejection fraction and CCI, along with higher (i.e. less negative) global longitudinal strain, all of which imply reduced LV contractility. Lower ERBB3 was also associated with higher LV mass. We then explored associations between plasma ERBB3 concentration and clinically used circulating biomarkers of cardiac status (**Table 4**). Lower ERBB3 was associated with higher concentrations of NT-proBNP (a marker of increased ventricular wall stress that is used in heart failure diagnosis) and troponin I (a marker of cardiomyocyte injury). These data suggest that lower circulating ERBB3, as seen in people with diabetes, is associated with impaired cardiac function, as assessed with routine clinical imaging and blood biomarkers.

**Table 4:**
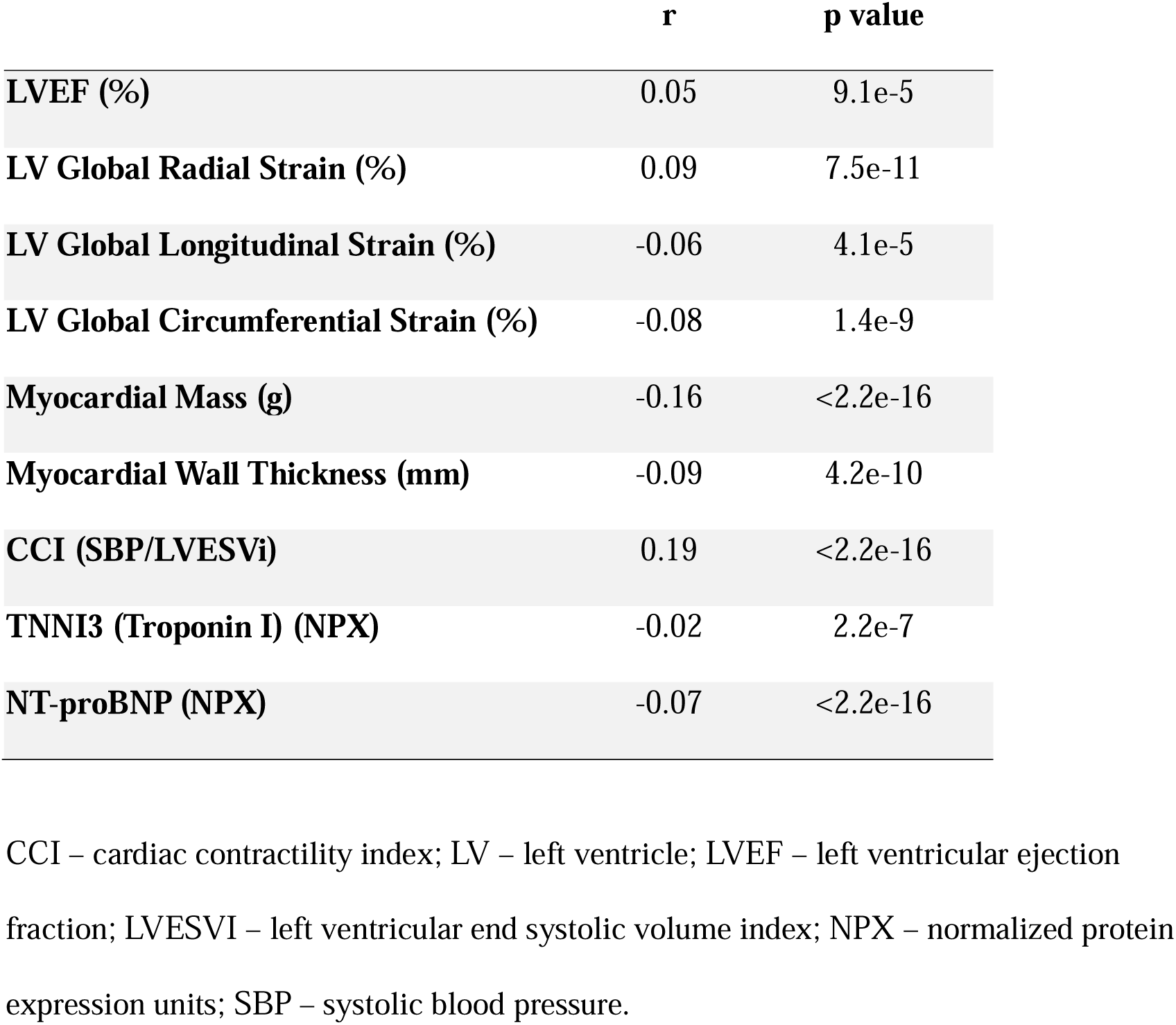
Correlation between ERBB3 and cardiovascular biomarkers in UKB.

|  | **r** | **p value** |
| --- | --- | --- |
| **LVEF (%)** | 0.05 | 9.1e-5 |
| **LV Global Radial Strain (%)** | 0.09 | 7.5e-11 |
| **LV Global Longitudinal Strain (%)** | -0.06 | 4.1e-5 |
| **LV Global Circumferential Strain (%)** | -0.08 | 1.4e-9 |
| **Myocardial Mass (g)** | -0.16 | <2.2e-16 |
| **Myocardial Wall Thickness (mm)** | -0.09 | 4.2e-10 |
| **CCI (SBP/LVESVi)** | 0.19 | <2.2e-16 |
| **TNNI3 (Troponin I) (NPX)** | -0.02 | 2.2e-7 |
| **NT-proBNP (NPX)** | -0.07 | <2.2e-16 |

CCI – cardiac contractility index; LV – left ventricle; LVEF – left ventricular ejection fraction; LVESVI – left ventricular end systolic volume index; NPX – normalized protein expression units; SBP – systolic blood pressure.

### 3.4 Cardiovascular outcomes of ERBB3 quartiles in UKB

We investigated whether low plasma ERBB3 protein is associated with long-term development of major cardiovascular events. During 719,262 person-years of follow-up, 2,234 incident heart failure events occurred. Kaplan-Meier curves illustrate a higher incidence of heart failure in participants in the lowest quartile of ERRB3, versus all other quartiles (**Figure 2A**). During 733,833 person-years of follow-up, 1,139 cardiovascular deaths occurred. Kaplan-Meier curves again illustrate a higher incidence of cardiovascular mortality in participants in the lowest quartile of ERRB3, versus all other quartiles (**Figure 2B**). Cox proportional hazards regression was then used to explore whether ERBB3 remained associated with these events after accounting for differing participant characteristics across ERBB3 quartiles (**Table 5**). After adjustment for age, sex, BMI, and DM, participants in the lowest quartile of plasma ERBB3 still exhibited greater risk of incident heart failure and cardiovascular mortality. As ERBB3 signals by heterodimerizing with other ERBB family members, we conducted similar analyses for ERBB2 and ERBB4 as UKB proteomic data are also available for these. We found similar, although less robust, patterns for both ERBB2 and ERBB4, with participants in the lowest quartile having higher risks of incident heart failure and cardiovascular mortality than some other quartiles when adjusted for age, sex, BMI and DM. (**Supplemental Figures 1-2** and **Supplemental Tables 6-7**). Finally, when adjusted for plasma NT-proBNP, participants in the lowest quartile of ERRB3 displayed greater risk of incident heart failure and cardiovascular mortality, versus all other quartiles (**Table 6**). These data suggest that ERBB3 is a circulating marker of major cardiovascular events, with additive value in relation to existing biomarkers.

**Figure 2:**
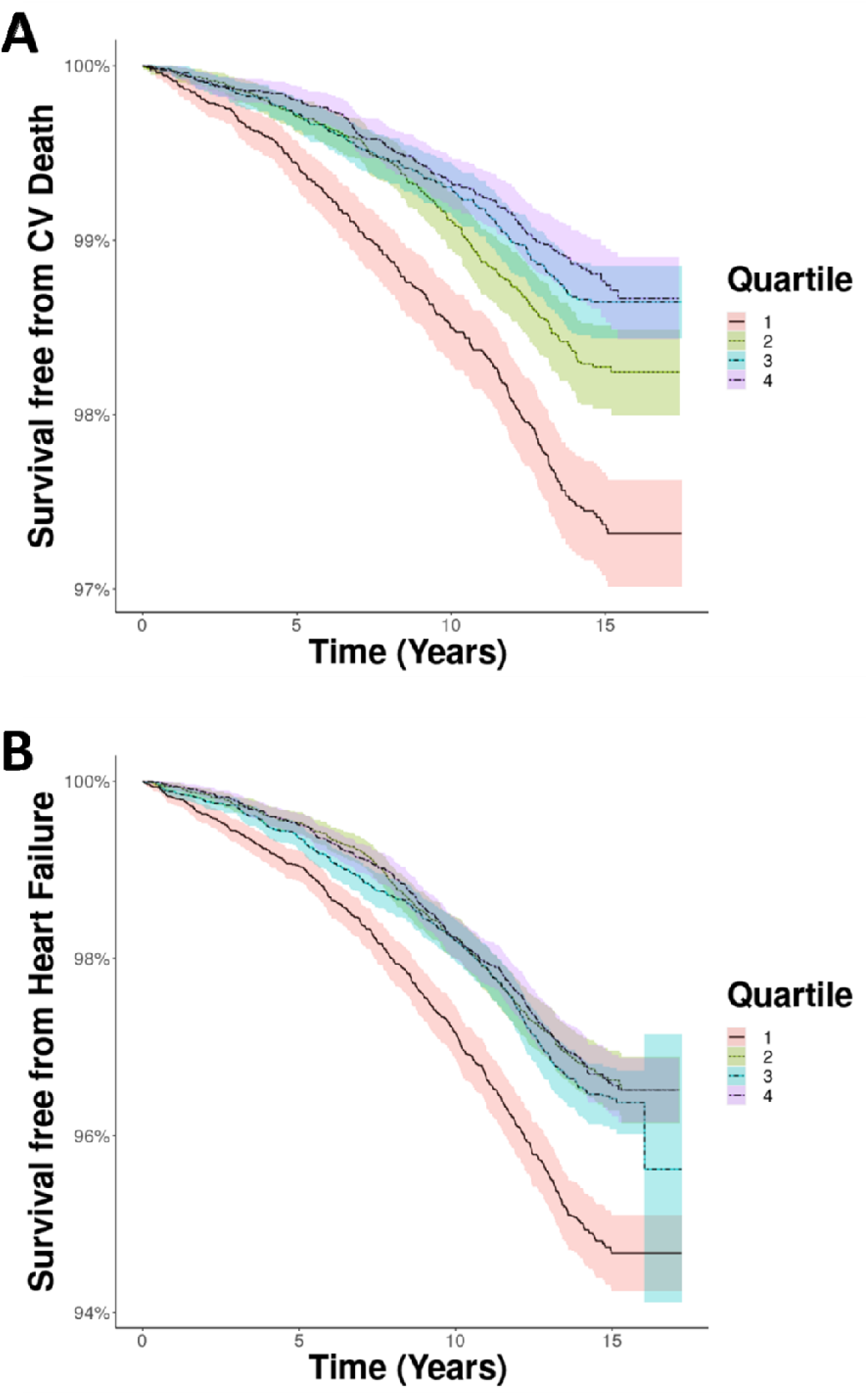
Incident heart failure and cardiovascular mortality in plasma ERBB3 quartiles. Kaplan-Meier curves illustrating probability of freedom from **A**) cardiovascular death and **B**) heart failure during long-term follow-up of the UKB cohort stratified into quartiles of plasma ERBB3, with 1 being the lowest quartile of expression and 4 the greatest.

**Table 5:** Adjusted risk of cardiovascular outcomes associated with ERBB3 quartiles.

|  | Cardiovascular Mortality |  | Heart Failure |  |
| --- | --- | --- | --- | --- |
|  | IRR (CI) | p value | IRR (CI) | p value |
| **ERBB3 Q1 vs Q2-4** | 1.24 (1.09-1.41) | 0.001 | 1.20 (1.09-1.31) | 0.000176 |
| **Age (per year)** | 1.11 (1.10-1.12) | <2e-16 | 1.12 (1.11-1.12) | <2e-16 |
| **Male sex** | 2.24 (1.95-2.57) | <2e-16 | 1.62 (1.48-1.77) | <2e-16 |
| **BMI (per Kg/m<sup>2</sup>)** | 1.06 (1.04-1.07) | <2e-16 | 1.08 (1.07-1.09) | <2e-16 |
| **DM** | 2.74 (2.34-3.20) | <2e-16 | 2.18 (1.94-2.45) | <2e-16 |

Q1 ERBB3 expression is the lowest quartile of expression, Q4 ERBB3 is the largest quartile of expression. IRR – incidence rate ratio; CI – 95% Confidence Interval; BMI – body mass index; DM – diabetes mellitus.

**Table 6:** Risk of cardiovascular outcomes associated with ERBB3 quartiles after adjusting for NT-proBNP.

|  | Cardiovascular Mortality |  | Heart Failure |  |
| --- | --- | --- | --- | --- |
|  | IRR (CI) | p value | IRR (CI) | p value |
| **ERBB3 Q1 vs Q2-4** | 1.48 (1.31-1.67) | 6.67e-10 | 1.34 (1.23 -1.45) | 1.38e-10 |
| **NT-proBNP**<br>**(per NPX unit)** | 1.77 (1.71-1.83) | <2e-16 | 1.75 (1.60-1.91) | <2e-16 |

Q1 ERBB3 expression is the lowest quartile of expression, Q4 ERBB3 is the largest quartile of expression. IRR – incidence rate ratio; CI – 95% Confidence Interval

## 4. Discussion

We set out to identify myocardial transcriptomic signatures associated with DM and then validate hits in their plasma protein form to identify potential biomarkers. The transcriptomic hits we identified in RA and LV myocardium did not overlap, suggesting distinct pathological processes in these chambers, although IgG immunoglobulin complexes were highlighted GO terms in both. Many of the DEGs and GO terms we identified have not previously been linked to diabetic heart disease and will be important to explore in future studies. In this analysis, we focused on the ERBB3 protein as the only myocardial DEG (detected in LV) to show directionally concordant differential expression as a plasma protein, being lower in people with DM. In the whole UKB plasma proteomics cohort, including participants without diabetes, we found lower circulating ERBB3 to be associated with impaired LV contractility (assessed with LVEF, GLS and CCI), incident heart failure and cardiovascular mortality. This is particularly interesting in the context of the cardiotoxicity known to arise from cancer therapies targeting ERBB3 and related family members. Notably, we also found some evidence that lower circulating ERBB2 and ERBB4 protein were associated with incident heart failure and cardiovascular mortality, supporting this receptor family as biomarkers of myocardial dysfunction. For ERBB3, we note that this is associated with adverse cardiovascular outcomes independent of the routine clinical biomarker NT-proBNP, suggesting a potential incremental value in detecting subclinical LV dysfunction.

Whilst many animal studies have set out to explore how experimentally induced DM impacts upon cardiac gene expression, few human studies have addressed this issue, largely because of the challenge of accessing tissue, especially from the LV.^22^ The largest published analysis of DM-associated LV gene expression profiles was conducted by Liu *et al*, who compared 7 people with type 2 DM and heart failure against 12 controls with only heart failure.^10^ They identified focal adhesions, vascular endothelial growth factor signalling, and mitogen-activated protein kinase signalling in pathway analyses. Our analysis did not highlight these pathways, but this may reflect the distinct focus of Liu *et al* on DM associated with heart failure. The larger sample size of our transcriptomic cohort and validation of ERBB3 using an alternate cohort and technology suggest our main finding is likely to be robust. The wider transcriptomic themes that we identified could not be studied using UKB plasma proteomic data and so require external validation using emerging transcriptomic cohorts or alternate approaches. In particular, a focus on immunoglobulin gene expression (presumably in B lymphocytes and plasma cells) is warranted, given enriched GO terms in RA and LV, along with emerging roles of B lymphocytes in heart.^23,24^ Moreover, our data suggest that separate studies of atrial and ventricular myocardium are essential, given that the transcriptomic signature of DM differed between these sites. Our data also suggest that human post mortem samples offer a valid route to biomarker and mechanism discovery, if technical factors are appropriately considered during analysis, potentially facilitating larger transcriptomic studies of LV myocardium which is difficult to acquire surgically.

Our finding that low circulating ERBB3 protein is associated with LV dysfunction and heart failure is further supported by data from patients receiving cancer therapeutics targeting ERBB2. Breast cancers commonly overexpress ERBB2 (also known as HER2), and the survival of people with these cancers is improved by agents, such as trastuzumab, that hinder epidermal growth factor signalling involving ERBB2.^25^ Notably, ERBB2 does not signal independently, but does so by heterodimerizing with other ERBB family members, including ERBB3^26^; indeed, the cancer therapeutic pertuzumab acts by hindering ERBB2-ERBB3 heterodimerization.^27^ Trastuzumab and pertuzumab are well known to increase the risk of cardiotoxicity, which manifests as reduced LV ejection fraction or even heart failure^28^; this risk is greater in people with DM.^29^ This observation led to the discovery that myocardial ERBB family receptors, including ERBB3, bind the ligand Neuregulin-1 (NRG1) that is released by cardiac endothelial cells in response to diverse stressors.^30^ Notably, recombinant forms of NRG1 have shown promise in augmenting LV contractility in heart failure and remain under active assessment in early phase clinical trials;^30,31^ they have also shown promise in a rat diabetic cardiomyopathy model.^32^ As with ERBB2, ERBB3 must heterodimerize to transmit intracellular signals, and does so with ERBB1, ERBB2 and ERBB4.^33^ Whilst not hits in myocardial RNA-seq analyses, our finding that low plasma ERBB2 and ERBB4 are also associated with incident heart failure and cardiovascular mortality adds further support to the importance of this interrelated family in myocardial dysfunction. Moreover, a recent multiomics study of human heart failure also implicated myocardial Erbb2 signaling^34^; our analysis extends this to people without heart failure, provides broader coverage of the ERBB receptor family, includes outcome data and shows the potential of ERBB members as circulating biomarkers.

The relationship between circulating ERBB3 protein and myocardial ERBB family signalling is unclear, although our data on ERBB3 (and ERBB2 and ERBB4) imply that lower circulating protein correspond to a phenotype expected from lower myocardial ERBB family signalling. The biological activity of circulating ERBB3 (e.g. as a decoy) is also unclear. *In vitro* studies suggest cleavage of ERBB3, amongst many other substrates, by the protease ADAM17 ^35^; the role of ERBB family cleavage in myocardial biology is poorly characterized and warrants more detailed study. However, myocardial ADAM17 knockout mice develop less myocardial dysfunction associated with diet-induced diabetes and it would be interesting to explore the role of ERBB3 in this phenotype.^36^ Irrespective of this uncertainty, our data suggest that circulating ERBB3 can add to existing heart failure biomarkers to detect people with early myocardial dysfunction and at risk of progression to heart failure. Whilst NT-proBNP is an excellent biomarker, our data suggest there is potential to use ERBB3 as a complementary marker in people with and without DM. It may also have a role in defining patients at risk of cardiotoxicity from HER2-targetted cancer therapy and in identifying potential responders to recombinant NRG1 in clinical trials.

Our study includes the largest described analysis of myocardial transcriptomic signatures associated with DM, followed by validation of hits in a very large plasma proteomic cohort that also allowed consideration of myocardial imaging and clinical outcomes for our principal hit. However, some limitations should be acknowledged. First, UKB proteomic data use Olink technology that quantifies proteins relative to the cohort mean concentration, rather than expressing these as absolute concentrations. This means that established cardiac biomarkers (Troponin I and NT-proBNP) and ERBB receptor family members have not been considered in light of clinically actionable thresholds. Hence, translation of our findings will warrant further work to cross-reference our data with other analytical approaches. It is also notable that this technology does not measure all circulating proteins, so is not truly unbiased. Second, our analyses could not stratify by type of DM due to the limitations of participant phenotyping (and also presumably statistical power). Therefore, whilst our findings apply to DM as a whole, they are probably heavily biased towards insights regarding type 2 DM. Since we explored UKB data in all comers (i.e. with and without diabetes), we expect that our findings will have broad relevance, but again we cannot make extensive subgroup analyses due to a lack of statistical power. Finally, our study is observational and so cannot directly infer a causal role of ERBB3 in myocardial dysfunction and adverse cardiovascular outcomes. Causal inference methods, such as Mendelian randomisation, could be used to extend our analyses. However, we argue that more compelling data for the ERBB receptor family directly contributing to myocardial (dys)function comes from the established cardiotoxicity of cancer therapies targeting these receptors and their signalling.

DM is associated with diverse and myocardial transcriptomic signatures, which are broadly distinct in the RA and LV, although altered IgG immunoglobin complex expression was noted in both sites. By assessing myocardial transcriptomic hits in their circulating protein form, we validated lower ERBB3 in people with diabetes and found this to be associated with LV dysfunction, incident heart failure and cardiovascular mortality. ERBB2 and ERBB4, closely related family members that heterodimerize with ERBB3, showed a similar pattern. These data suggest that ERBB signalling may be important in the development of heart failure in people with diabetes, and ERBB3 could represent a useful biomarker to identify this risk and target preventative therapies.

## Data Availability

All data produced are available online at https://www.ukbiobank.ac.uk and https://gtexportal.org/

https://gtexportal.org/

## Acknowledgements

This research has been conducted using the UK Biobank Resource under Application Number 105351. This work uses data provided by patients and collected by the NHS as part of their care and support; Copyright © (2022), NHS Digital; Re-used with the permission of UK Biobank. All rights reserved. This research used data assets made available by National Safe Haven as part of the Data and Connectivity National Core Study, led by Health Data Research UK in partnership with the Office for National Statistics and funded by UK Research and Innovation (research which commenced between 1st October 2020 – 31st March 2021 grant ref MC_PC_20029; 1st April 2021-30th September 2022 grant ref MC_PC_20058). The Genotype-Tissue Expression (GTEx) Project was supported by the Common Fund of the Office of the Director of the National Institutes of Health, and by NCI, NHGRI, NHLBI, NIDA, NIMH, and NINDS. The data used for the analyses described in this manuscript were obtained from: the GTEx Portal on 08/22/2022 and/or dbGaP accession number phs000424.v8.p2. CC is supported by BHF Mautner Career Development Fellowship. EL is funded by Wellcome Trust Clinical Career Development Fellowship (221690/Z/20/Z) and receives support from National Institute for Health and Care Research (NIHR) Leeds Biomedical Research Centre. The views expressed are those of the author(s) and not necessarily those of the NHS, the NIHR or the Department of Health and Social Care.

## Author Contribution Statement

MCR performed computational analyses. MCR and RC drafted the manuscript. All authors conceptualised the study and contributed to the analysis plan. All authors revised and approved the final manuscript.

