## Supplemental for "The diabetic myocardial transcriptome reveals Erbb3 as a novel biomarker of incident heart failure"

**Supplementary Table 1: Differentially expressed genes in DM vs non-DM left ventricle using GTEx data.**

| GENE ENSEMBL ID | Base Mean | L2FC | lfcSE | padj | GENE NAME |
| --- | --- | --- | --- | --- | --- |
| ENSG00000225972.1 | 9256.7 | -1.227 | 0.271 | 2.3E-04 | MTND1P23 |
| ENSG00000102385.12 | 25.6 | -0.756 | 0.185 | 1.6E-03 | DRP2 |
| ENSG00000206579.8 | 100.7 | -0.684 | 0.149 | 2.3E-04 | XKR4 |
| ENSG00000237515.8 | 25.2 | -0.647 | 0.132 | 1.6E-04 | SHISA9 |
| ENSG00000147588.6 | 57.7 | -0.583 | 0.158 | 7.2E-03 | PMP2 |
| ENSG00000115665.8 | 16.3 | -0.572 | 0.208 | 3.9E-02 | SLC5A7 |
| ENSG00000158887.15 | 303.0 | -0.546 | 0.111 | 1.6E-04 | MPZ |
| ENSG00000110900.14 | 378.6 | -0.531 | 0.112 | 2.3E-04 | TSPAN11 |
| ENSG00000026559.13 | 28.6 | -0.507 | 0.139 | 8.7E-03 | KCNG1 |
| ENSG00000163873.9 | 89.1 | -0.502 | 0.140 | 6.6E-03 | GRIK3 |
| ENSG00000215018.9 | 266.5 | -0.500 | 0.128 | 3.4E-03 | COL28A1 |
| ENSG00000162951.10 | 26.1 | -0.478 | 0.154 | 2.5E-02 | LRRTM1 |
| ENSG00000160307.9 | 152.4 | -0.468 | 0.135 | 8.6E-03 | S100B |
| ENSG00000183960.8 | 20.3 | -0.462 | 0.131 | 7.8E-03 | KCNH8 |
| ENSG00000123560.13 | 525.9 | -0.461 | 0.118 | 3.5E-03 | PLP1 |
| ENSG00000125851.9 | 108.6 | -0.455 | 0.142 | 1.7E-02 | PCSK2 |
| ENSG00000065361.14 | 290.5 | -0.448 | 0.100 | 3.7E-04 | ERBB3 |
| ENSG00000179915.22 | 172.1 | -0.433 | 0.114 | 3.5E-03 | NRXN1 |
| ENSG00000223403.3 | 97.7 | -0.431 | 0.118 | 7.8E-03 | MEG9 |
| ENSG00000198910.12 | 545.8 | -0.412 | 0.102 | 3.2E-03 | L1CAM |
| ENSG00000185985.8 | 60.9 | -0.411 | 0.131 | 2.1E-02 | SLITRK2 |
| ENSG00000092758.15 | 296.8 | -0.381 | 0.105 | 7.2E-03 | COL9A3 |
| ENSG00000021645.18 | 107.5 | -0.377 | 0.114 | 1.5E-02 | NRXN3 |
| ENSG00000105855.9 | 250.0 | -0.366 | 0.120 | 2.7E-02 | ITGB8 |
| ENSG00000108018.15 | 240.7 | -0.348 | 0.109 | 2.3E-02 | SORCS1 |
| ENSG00000272941.1 | 91.9 | 0.331 | 0.098 | 1.7E-02 | RP11-134L10.1 |
| ENSG00000126803.9 | 1107.6 | 0.353 | 0.113 | 3.4E-02 | HSPA2 |
| ENSG00000275457.1 | 23.2 | 0.359 | 0.122 | 4.1E-02 | RP1-198K11.5 |
| ENSG00000006555.10 | 17.1 | 0.507 | 0.159 | 1.7E-02 | TTC22 |
| ENSG00000179292.4 | 8.1 | 0.637 | 0.197 | 1.7E-02 | TMEM151A |
| ENSG00000158050.4 | 483.5 | 0.670 | 0.186 | 5.5E-03 | DUSP2 |
| ENSG00000211892.3 | 59.1 | 1.381 | 0.356 | 1.6E-04 | IGHG4 |

Differential expression is defined as pAdj <0.05 and L2FC >0.32 or <-0.32. L2FC - Log2 Fold Change; LFCSE - Log2 Fold Change Standard Error.

**Supplementary Table 2: Differentially expressed genes in DM vs non-DM atrial appendage using GTEx data. L2FC = Log_2_ Fold Change. Differential expression is defined as p adj <0.05 and LFC >0.32 or <-0.32**

| GENE ENSEMBL ID | Base Mean | L2FC | lfcSE | padj | GENE NAME |
| --- | --- | --- | --- | --- | --- |
| ENSG00000211679.2 | 125.5 | -1.152 | 0.272 | 5.3E-04 | IGLC3 |
| ENSG00000120907.17 | 40.7 | -0.960 | 0.209 | 2.6E-04 | ADRA1A |
| ENSG00000211895.4 | 897.9 | -0.919 | 0.225 | 1.7E-03 | IGHA1 |
| ENSG00000184956.15 | 29.5 | -0.794 | 0.206 | 6.3E-03 | MUC6 |
| ENSG00000170476.15 | 27.5 | -0.780 | 0.227 | 1.0E-02 | MZB1 |
| ENSG00000263961.6 | 13.4 | -0.608 | 0.171 | 1.4E-02 | C1orf186 |
| ENSG00000026751.16 | 38.8 | -0.591 | 0.160 | 8.6E-03 | SLAMF7 |
| ENSG00000121807.5 | 44.9 | -0.505 | 0.146 | 2.1E-02 | CCR2 |
| ENSG00000265206.5 | 36.5 | -0.493 | 0.152 | 2.5E-02 | RP5-1171I10.5 |
| ENSG00000169851.15 | 249.4 | -0.468 | 0.154 | 3.0E-02 | PCDH7 |
| ENSG00000100033.16 | 226.5 | -0.463 | 0.128 | 1.1E-02 | PRODH |
| ENSG00000075702.16 | 1105.0 | -0.455 | 0.134 | 1.6E-02 | WDR62 |
| ENSG00000258498.8 | 444.9 | -0.409 | 0.149 | 4.3E-02 | DIO3OS |
| ENSG00000281969.1 | 11.6 | -0.409 | 0.135 | 3.4E-02 | XXyac-YR38GF2.1 |
| ENSG00000197977.3 | 284.8 | -0.394 | 0.109 | 1.1E-02 | ELOVL2 |
| ENSG00000173198.5 | 51.4 | -0.385 | 0.120 | 2.4E-02 | CYSLTR1 |
| ENSG00000161267.11 | 1110.5 | -0.382 | 0.113 | 1.6E-02 | BDH1 |
| ENSG00000103175.10 | 138.4 | -0.376 | 0.132 | 4.3E-02 | WFDC1 |
| ENSG00000140090.17 | 96.8 | -0.368 | 0.104 | 1.4E-02 | SLC24A4 |
| ENSG00000167244.18 | 7471.3 | -0.358 | 0.105 | 1.4E-02 | IGF2 |
| ENSG00000181234.9 | 730.5 | -0.357 | 0.117 | 3.5E-02 | TMEM132C |
| ENSG00000106789.12 | 43.9 | -0.355 | 0.097 | 1.1E-02 | CORO2A |
| ENSG00000166501.12 | 185.3 | -0.346 | 0.104 | 2.1E-02 | PRKCB |
| ENSG00000122824.10 | 65.7 | -0.343 | 0.112 | 3.1E-02 | NUDT10 |
| ENSG00000135074.15 | 4696.4 | 0.376 | 0.125 | 3.1E-02 | ADAM19 |
| ENSG00000268894.6 | 18.9 | 0.401 | 0.132 | 2.5E-02 | PLCE1-AS1 |
| ENSG00000266954.1 | 28.5 | 0.434 | 0.161 | 4.7E-02 | RP11-701H16.4 |
| ENSG00000152315.4 | 137.4 | 0.476 | 0.145 | 2.3E-02 | KCNK13 |
| ENSG00000140519.12 | 108.3 | 0.553 | 0.177 | 2.1E-02 | RHCG |
| ENSG00000181577.15 | 155.2 | 0.570 | 0.227 | 3.5E-02 | C6orf223 |
| ENSG00000278022.1 | 13.5 | 0.674 | 0.232 | 3.4E-02 | RP11-35O15.2 |
| ENSG00000135346.8 | 42.9 | 0.887 | 0.280 | 2.3E-02 | CGA |

Differential expression is defined as pAdj <0.05 and L2FC >0.32 or <-0.32. L2FC - Log2 Fold Change; LFCSE - Log2 Fold Change Standard Error.

**Supplemental Table 3: LV gProfiler results of 32 LV DEGs from GTEx analysis**

| **Source** | **Term Name** | **Term ID** | **pAdj** | **Term size** | | **Query size** | | **Intersection size** | | **Intersection Genes** |
| --- | --- | --- | --- | --- | --- | --- | --- | --- | --- | --- |
| GO:MF | neuroligin family protein binding | GO:0097109 | 3.E-03 | 5 | 25 | | 2 | | NRXN1,NRXN3 | |
| GO:MF | calcium channel regulator activity | GO:0005246 | 4.E-02 | 45 | 25 | | 2 | | NRXN1,NRXN3 | |
| GO:MF | extracellular matrix structural constituent conferring tensile strength | GO:0030020 | 4.E-02 | 41 | 25 | | 2 | | COL28A1,COL9A3 | |
| GO:MF | choline:sodium symporter activity | GO:0005307 | 4.E-02 | 1 | 25 | | 1 | | SLC5A7 | |
| GO:MF | potassium channel activity | GO:0005267 | 4.E-02 | 124 | 25 | | 3 | | KCNG1,GRIK3,KCNH8 | |
| GO:MF | tau protein binding | GO:0048156 | 4.E-02 | 43 | 25 | | 2 | | S100B,HSPA2 | |
| GO:BP | synaptic signaling | GO:0099536 | 1.E-05 | 778 | 26 | | 10 | | DRP2,SHISA9,SLC5A7,  MPZ,GRIK3,LRRTM1,  S100B,PLP1,NRXN1,  NRXN3 | |
| GO:BP | axon development | GO:0061564 | 3.E-03 | 509 | 26 | | 6 | | S100B,PLP1,NRXN1,  L1CAM,SLITRK2,NRXN3 | |
| GO:BP | vocalization behavior | GO:0071625 | 1.E-02 | 19 | 26 | | 2 | | NRXN1,NRXN3 | |
| GO:BP | ionotropic glutamate receptor signaling pathway | GO:0035235 | 1.E-02 | 24 | 26 | | 2 | | GRIK3,PLP1 | |
| GO:BP | synaptic membrane adhesion | GO:0099560 | 1.E-02 | 29 | 26 | | 2 | | NRXN1,SLITRK2 | |
| GO:BP | islet amyloid polypeptide processing | GO:0034231 | 2.E-02 | 1 | 26 | | 1 | | PCSK2 | |
| GO:BP | protein-containing complex assembly involved in synapse maturation | GO:0090126 | 2.E-02 | 1 | 26 | | 1 | | NRXN1 | |
| GO:BP | social behavior | GO:0035176 | 2.E-02 | 53 | 26 | | 2 | | NRXN1,NRXN3 | |
| GO:BP | guanylate kinase-associated protein clustering | GO:0097117 | 3.E-02 | 2 | 26 | | 1 | | NRXN1 | |
| GO:BP | synaptonemal complex disassembly | GO:0070194 | 3.E-02 | 2 | 26 | | 1 | | HSPA2 | |
| GO:BP | enkephalin processing | GO:0034230 | 3.E-02 | 2 | 26 | | 1 | | PCSK2 | |
| GO:BP | regulation of monoatomic ion transmembrane transport | GO:0034765 | 3.E-02 | 467 | 26 | | 4 | | KCNG1,KCNH8,PLP1,  HSPA2 | |
| GO:BP | positive regulation of cardiac muscle tissue development | GO:0055025 | 3.E-02 | 2 | 26 | | 1 | | ERBB3 | |
| GO:BP | receptor localization to synapse | GO:0097120 | 3.E-02 | 64 | 26 | | 2 | | NRXN1,NRXN3 | |
| GO:BP | acetylcholine biosynthetic process | GO:0008292 | 3.E-02 | 3 | 26 | | 1 | | SLC5A7 | |
| GO:BP | neuroligin clustering involved in postsynaptic membrane assembly | GO:0097118 | 3.E-02 | 3 | 26 | | 1 | | NRXN1 | |
| GO:BP | metal ion transport | GO:0030001 | 4.E-02 | 903 | 26 | | 5 | | SLC5A7,KCNG1,KCNH8,  PLP1,HSPA2 | |
| GO:BP | learning or memory | GO:0007611 | 4.E-02 | 275 | 26 | | 3 | | S100B,NRXN1,NRXN3 | |
| GO:BP | ERBB2-ERBB3 signaling pathway | GO:0038133 | 4.E-02 | 4 | 26 | | 1 | | ERBB3 | |
| GO:BP | Langerhans cell differentiation | GO:0061520 | 4.E-02 | 4 | 26 | | 1 | | ITGB8 | |
| GO:BP | sympathetic neuron projection extension | GO:0097490 | 4.E-02 | 5 | 26 | | 1 | | S100B | |
| GO:BP | vocal learning | GO:0042297 | 4.E-02 | 5 | 26 | | 1 | | NRXN1 | |
| GO:BP | positive regulation of calcium ion transmembrane transport | GO:1904427 | 4.E-02 | 93 | 26 | | 2 | | PLP1,HSPA2 | |
| GO:BP | gamma-aminobutyric acid receptor clustering | GO:0097112 | 4.E-02 | 5 | 26 | | 1 | | NRXN1 | |
| GO:BP | NMDA glutamate receptor clustering | GO:0097114 | 4.E-02 | 5 | 26 | | 1 | | NRXN1 | |
| GO:BP | positive regulation of ATPase-coupled calcium transmembrane transporter activity | GO:1901896 | 5.E-02 | 6 | 26 | | 1 | | HSPA2 | |
| GO:BP | negative regulation of filopodium assembly | GO:0051490 | 5.E-02 | 6 | 26 | | 1 | | NRXN1 | |
| GO:BP | regulation of grooming behavior | GO:2000821 | 5.E-02 | 6 | 26 | | 1 | | NRXN1 | |
| GO:CC | neuronal cell body | GO:0043025 | 2.E-04 | 500 | 28 | | 7 | | DRP2,SLC5A7,GRIK3,  S100B,PCSK2,NRXN1,  L1CAM | |
| GO:CC | cell periphery | GO:0071944 | 2.E-04 | 6202 | 28 | | 20 | | DRP2,XKR4,SHISA9,  SLC5A7,MPZ,KCNG1,  GRIK3,COL28A1,LRRTM1,  KCNH8,PLP1,ERBB3,NRXN1,  L1CAM,SLITRK2,COL9A3,  NRXN3,ITGB8,HSPA2,  IGHG4 | |
| GO:CC | myelin sheath | GO:0043209 | 4.E-04 | 45 | 28 | | 3 | | PMP2,MPZ,PLP1 | |
| GO:CC | collagen type V trimer | GO:0005588 | 2.E-02 | 4 | 28 | | 1 | | COL28A1 | |
| GO:CC | collagen type IX trimer | GO:0005594 | 2.E-02 | 4 | 28 | | 1 | | COL9A3 | |
| GO:CC | extracellular space | GO:0005615 | 2.E-02 | 3284 | 28 | | 10 | | PMP2,COL28A1,LRRTM1,  S100B,PCSK2,ERBB3,  COL9A3,ITGB8,HSPA2,  IGHG4 | |
| GO:CC | IgG immunoglobulin complex | GO:0071735 | 2.E-02 | 5 | 28 | | 1 | | IGHG4 | |
| GO:CC | meiotic spindle | GO:0072687 | 5.E-02 | 15 | 28 | | 1 | | HSPA2 | |
| GO:CC | CatSper complex | GO:0036128 | 5.E-02 | 15 | 28 | | 1 | | HSPA2 | |

**Supplemental Table 4: gProfiler results of 32 RA DEGs from the GTEx analysis**

| Source | Term name | Term id | pAdj | Term size | Query size | Intersection size | Intersection genes |
| --- | --- | --- | --- | --- | --- | --- | --- |
| GO:BP | B cell receptor signaling pathway | GO:0050853 | 0.044417 | 70 | 24 | 3 | IGLC3,IGHA1,PRKCB |
| GO:BP | regulation of insulin receptor signaling pathway | GO:0046626 | 0.044417 | 67 | 24 | 3 | MZB1,IGF2,PRKCB |
| GO:CC | IgG immunoglobulin complex | GO:0071735 | 0.001235 | 5 | 24 | 2 | IGLC3,IGHA1 |
| GO:CC | IgA immunoglobulin complex | GO:0071745 | 0.001235 | 6 | 24 | 2 | IGLC3,IGHA1 |
| GO:CC | side of membrane | GO:0098552 | 0.049962 | 656 | 24 | 4 | IGLC3,SLAMF7,CCR2,  BDH1 |
| GO:CC | IgM immunoglobulin complex | GO:0071753 | 0.049962 | 4 | 24 | 1 | IGLC3 |
| GO:CC | Golgi lumen | GO:0005796 | 0.049962 | 103 | 24 | 2 | MUC6,CGA |
| GO:CC | IgE immunoglobulin complex | GO:0071742 | 0.049962 | 2 | 24 | 1 | IGLC3 |
| GO:CC | IgD immunoglobulin complex | GO:0071738 | 0.049962 | 1 | 24 | 1 | IGLC3 |
| GO:CC | platelet alpha granule | GO:0031091 | 0.049962 | 90 | 24 | 2 | PCDH7,IGF2 |
| GO:CC | follicle-stimulating hormone complex | GO:0016914 | 0.049962 | 2 | 24 | 1 | CGA |
| GO:CC | plasma membrane | GO:0005886 | 0.049962 | 5717 | 24 | 13 | IGLC3,ADRA1A,IGHA1,  MUC6,SLAMF7,CCR2,PCDH7,CYSLTR1,SLC24A4,PRKCB,ADAM19,KCNK13,RHCG |

**Supplemental Table 5: Adjusted risk of cardiovascular outcomes associated with ERBB2 quartiles**

|  | **Cardiovascular Mortality** | | **Heart Failure** | |
| --- | --- | --- | --- | --- |
|  | **IRR (CI)** | **p** | **IRR (CI)** | **p** |
| **ERBB2** |  |  |  |  |
| **Q1 vs Q2** | 1.21 (1.01-1.45) | 0.0414 | 1.16 (1.07-1.26) | 0.0267 |
| **Q1 vs Q3** | 1.21 (1.01-1.45) | 0.0364 | 1.08 (0.98-1.18) | 0.250 |
| **Q1 vs Q4** | 1.01 (0.85-1.20) | 0.9096 | 0.95 (0.87-1.04) | 0.455 |
| **Age (per year)** | 1.11 (1.10-1.13) | <2e-16 | 1.12 (1.11-1.13) | <2e-16 |
| **Male sex** | 2.38 (2.09-2.71) | <2e-16 | 1.69 (1.51-1.90) | <2e-16 |
| **BMI (per Kg/m^2^)** | 1.05 (1.04-1.07) | <2e-16 | 1.08 (1.00-1.16) | <2e-16 |
| **DM** | 2.72 (2.33-3.18) | <2e-16 | 2.21 (2.06-2.31) | <2e-16 |

Q1 ERBB2 expression is the lowest quartile of expression, Q4 ERBB2 is the largest quartile of expression. IRR – incidence rate ratio; CI – 95% Confidence Interval; BMI – body mass index; DM – diabetes mellitus.

**Supplemental Table 6:** **Adjusted risk of cardiovascular outcomes associated with ERBB4 quartiles**

|  | **Cardiovascular Mortality** | | **Heart Failure** | |
| --- | --- | --- | --- | --- |
|  | **IRR (CI)** | **p** | **IRR (CI)** | **p** |
| **ERBB4** |  |  |  |  |
| **Q1 vs Q2** | 1.19 (1.00-1.41) | 0.049 | 1.31 (1.17-1.48) | 0.0267 |
| **Q1 vs Q3** | 1.05 (0.89-1.24) | 0.581 | 1.23 (1.09-1.38) | 0.250 |
| **Q1 vs Q4** | 0.95 (0.81-1.13) | 0.573 | 1.06 (0.95-1.19) | 0.455 |
| **Age (per year)** | 1.11 (1.10-1.12) | <2e-16 | 1.12 (1.11-1.13) | <2e-16 |
| **Male sex** | 2.41 (2.11-2.74) | <2e-16 | 1.71 (1.57-1.86) | <2e-16 |
| **BMI (per Kg/m^2^)** | 1.06 (1.04-1.07) | <2e-16 | 1.08 (1.07-1.08) | <2e-16 |
| **DM** | 2.70 (2.31-3.16) | <2e-16 | 2.20 (1.96-2.47) | <2e-16 |

Q1 ERBB4 expression is the lowest quartile of expression, Q4 ERBB4 is the largest quartile of expression. IRR – incidence rate ratio; CI – 95% Confidence Interval; BMI – body mass index; DM – diabetes mellitus.

**Supplemental Figure 1: Incident heart failure and cardiovascular mortality in plasma ERBB2 quartiles**

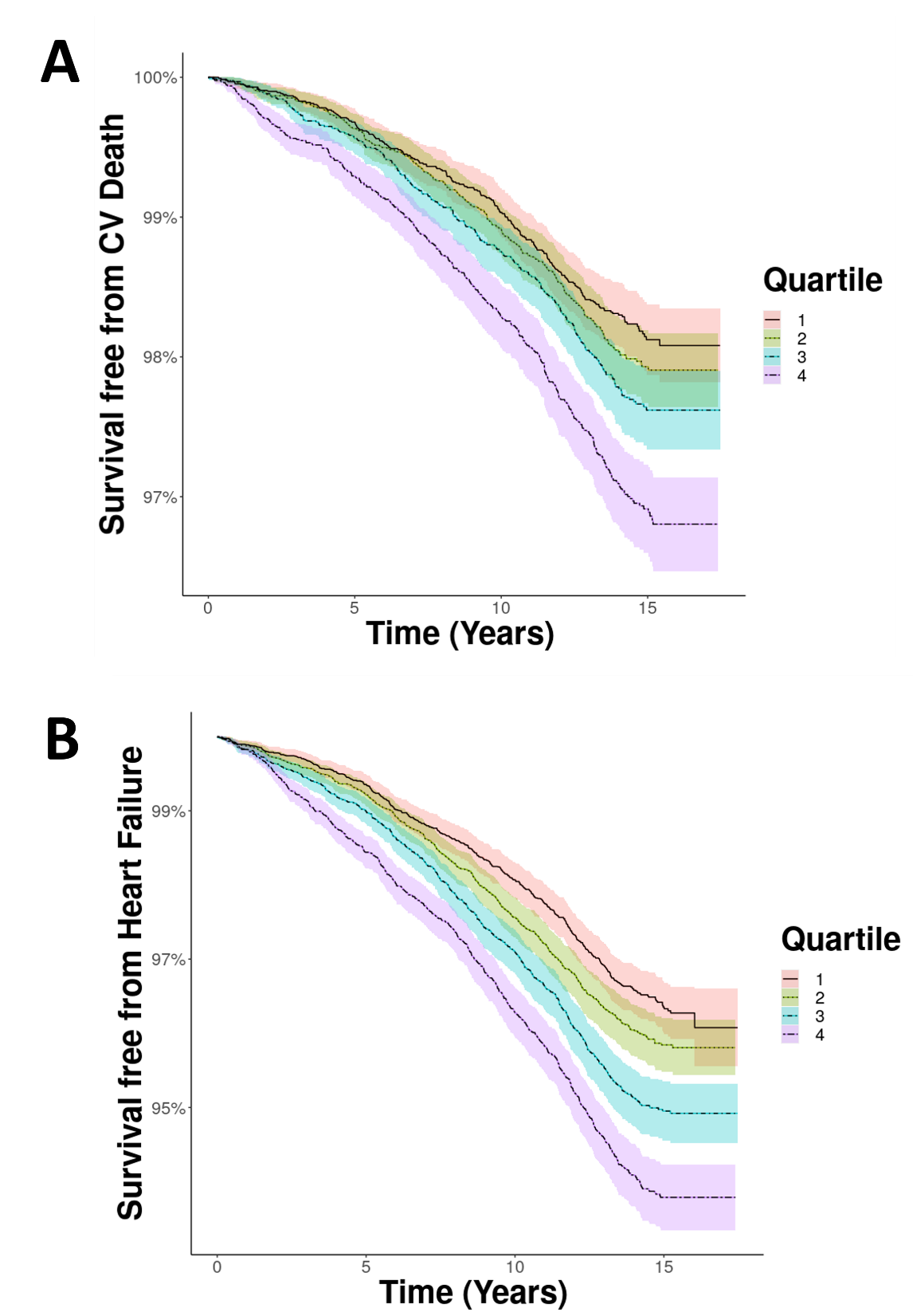

Kaplan-Meier curves illustrating probability of freedom from **A**) cardiovascular death and **B**) heart failure during long-term follow-up of the UKB cohort stratified into quartiles of plasma ERBB2, with 1 being the lowest quartile of expression and 4 the greatest.

**Supplemental Figure 2: Incident heart failure and cardiovascular mortality in plasma ERBB4 quartiles**

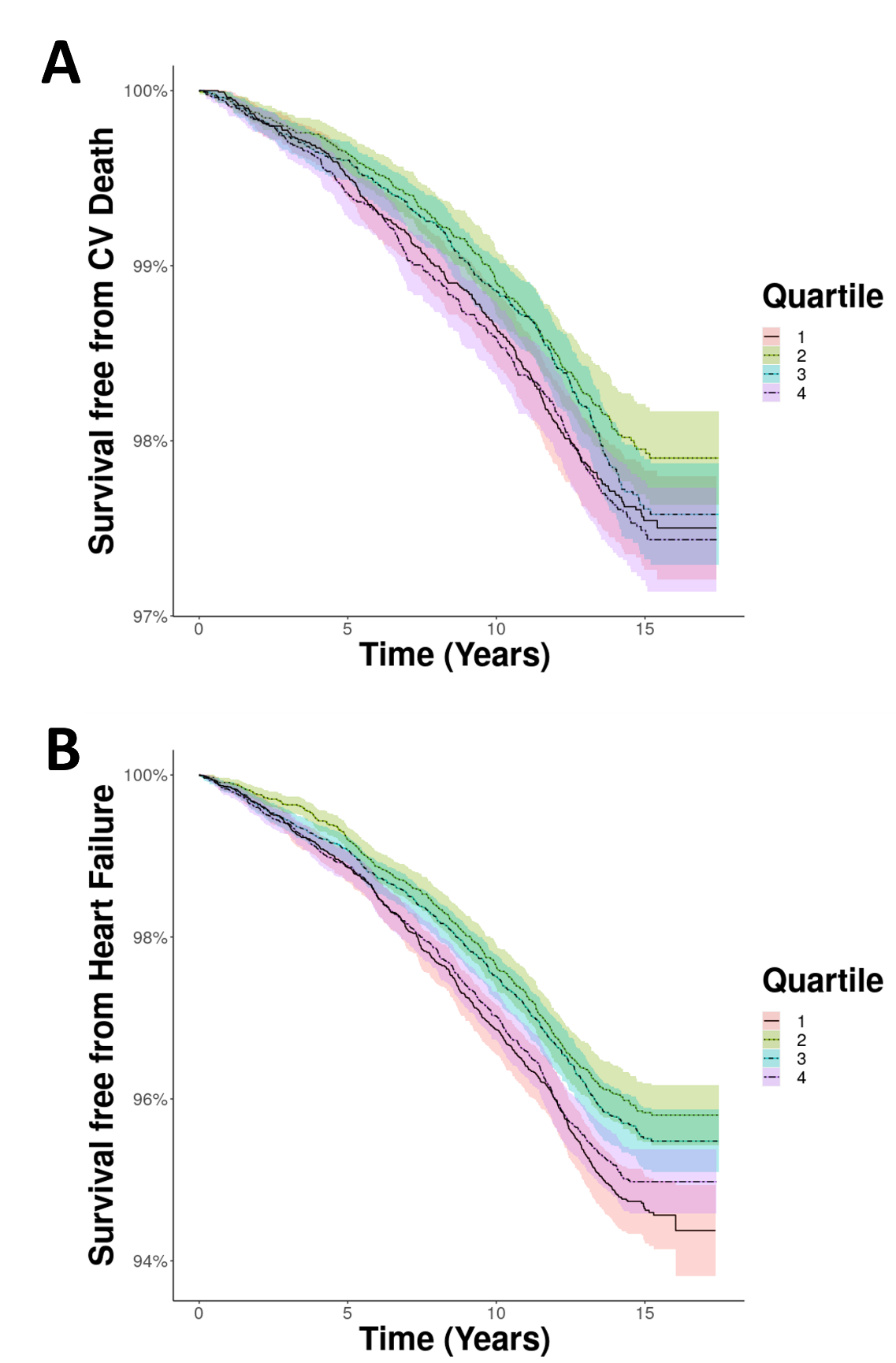

Kaplan-Meier curves illustrating probability of freedom from **A**) cardiovascular death and **B**) heart failure during long-term follow-up of the UKB cohort stratified into quartiles of plasma ERBB4, with 1 being the lowest quartile of expression and 4 the greatest.
